# CNVscore calculates pathogenicity scores for copy number variants together with uncertainty estimates accounting for learning biases in reference Mendelian disorder datasets

**DOI:** 10.1101/2022.06.23.22276396

**Authors:** Francisco Requena, David Salgado, Valérie Malan, Damien Sanlaville, Frédéric Bilan, Christophe Béroud, Antonio Rausell

## Abstract

Copy number variants (CNVs) are a major cause of rare pediatric diseases with a broad spectrum of phenotypes. Genetic diagnosis based on comparative genomic hybridization tests typically identifies ∼8-10% of patients as having CNVs of unknown significance, revealing the current limits of clinical interpretation. The adoption of whole-genome sequencing (WGS) as a first-line genetic test has significantly increased the load of CNVs identified in single genomes. Alongside short- and long-read sequencing technologies, a number of pathogenicity scores have been developed for filtering and prioritizing large sets of candidate CNVs in clinical settings. However, current approaches are often based, either explicitly or implicitly, on clinically annotated reference sets, which are likely to bias their predictions. In this study we developed CNVscore, a supervised-learning approach combining tree ensembles and a Bayesian classifier trained on pathogenic and non-pathogenic CNVs from reference databases. Unlike previous approaches, CNVscore couples pathogenicity estimates with uncertainty scores, making it possible to evaluate the suitability of a model for the query CNVs. Comprehensive comparative benchmark tests across independent sets and against alternative methods showed that CNVscore effectively distinguishes between pathogenic and benign CNVs. We also found that CNVs associated with CNVscores of low uncertainty were predicted with significantly higher accuracy than those of high uncertainty. However, the performance of current scoring approaches, including CNVscore, was compromised on CNV sets enriched in highly uncertain variants and presenting unconventional features, such as functionally relevant non-coding elements or the presence of disease genes irrelevant for the clinical phenotypes investigated. Finally, we used the CNVscore framework to guide CNV scoring model selection for the French National Database of Constitutional CNVs (BANCCO), which includes clinical diagnosis annotations. The CNVscore framework provides an objective strategy for leveraging the uncertainty on bioinformatic predictions to enhance the assessment of CNV pathogenicity in rare-disease cohorts. CNVscore is available as open-source software from https://github.com/RausellLab/CNVscore and is integrated into the CNVxplorer webserver http://cnvxplorer.com.

## Introduction

Copy number variants (CNVs) are a major cause of rare genetic diseases. The most widely adopted clinical cytogenetic tests for CNVs are currently chromosomal microarray tests (CMAs), such as array comparative genomic hybridization (aCGH) and single-nucleotide polymorphism (SNP) arrays, which are performed on DNA from whole blood, tissue or amniotic fluid. CMAs have a genomic resolution for routine diagnosis of ∼10–300 kilobases, resulting in the identification of small numbers of candidate CNV events per patient, typically between 0 and 10, depending on the analysis settings. This low throughput has remained compatible with clinical assessments based on expert curation. However, whole-genome sequencing (WGS) is increasingly being adopted as a first-line genetic test in genomic medicine [1]. One of the major advantages of WGS over whole-exome sequencing (WES) is that WGS allows the simultaneous and accurate genome-wide calling of single-nucleotide variants (SNVs), CNVs and other types of structural variants (SVs) [2]. Moreover, short-read WGS-based CNV calling identifies a larger number of events of smaller size than CMAs. Indeed, recent large-scale genome sequencing projects have estimated a median of 3,505 deletions and 723 duplications per genome, with a median length of 756 base pairs (bp) and 4,283 bp respectively (gnomAD-SV GnomAD)[1]. From the clinical diagnosis standpoint, one of the major practical consequences of the adoption of WGS is an increase in the number of candidate CNVs to be evaluated. A second consequence is that the characteristics of such candidate variants are different, with a higher proportion having no direct impact on protein-coding gene, due to a significant shift towards the detection of shorter CNVs.

Various bioinformatics webtools [3,4] and pathogenicity scores (**Table 1**) have recently been developed to facilitate the annotation, filtering and clinical assessment of large numbers of candidate CNVs in patients with rare diseases. Two major categories of scores can be distinguished: (a) those implementing heuristics based on reference medical guidelines, such as ACMG and ClinGen [4] and (b) those based on supervised-learning approaches trained on reference pathogenic and non-pathogenic CNV datasets (**Table 1**). Despite their methodological differences, both these approaches are highly dependent on the reference genetic variants (SV and SNVs) and their clinical consequences reported at a given time, resulting in a risk of bias in their predictions. Thus, while the influence of prior knowledge on predictive outcome is obvious in the case of medical guidelines, it is often overlooked in the application of supervised-learning approaches. There are two types of methods: (i) those explicitly considering known clinical annotations of the targeted genomic elements (for instance, gene-disease associations) as predictive features, and (ii) those based purely on *unbiased* features throughout the genome, such as sequence conservation or epigenetic marks (**Table 1**). However, even this second group of methods may be prone to severe bias in their predictions for query CNVs partially overlapping the CNV sets used in the training process.

**Table 1.** State-of-the-art CNV scoring methods. This table summarizes the major characteristics of four supervised learning-based tools (STRVCTVRE, TADA, CADD-SV and X-CNV) and one guideline-based approach (ClassifyCNV). The features used by supervised-learning approaches were classified into two major categories: genome-wide features (purportedly unbiased) and knowledge-based features (conveying prior evidence about the disease-association characteristics of genomic elements within the CNV boundaries). Two features were identified for knowledge-based features: HI index, a genome-wide score based on a list of known human haploinsufficient (HI) genes; and the Developmental Disorders Genotype-to-Phenotype database (DDG2P) gene list [9]. Asterisks (*) identify two tools (TADA and X-CNV) presenting potential contamination between their training datasets and the benchmark datasets used in this work, as no prevention strategy for downstream applications was used for these methods. Thus, TADA included pathogenic CNVs from Decipher and non-pathogenic CNVs from gnomAD and Audano et al., 2019. X-CNV used the dbVar database for training, which includes CNVs from Clinvar, gnomAD and Audano et al, 2019.

| Name | Type of feature | Knowledge-based features | Genome assembly | Learning method | Positive obs. sources (Training data) | Negative obs. sources (Training data) | Length-matched training set | Independent datasets (held-out test) | Ref |
| --- | --- | --- | --- | --- | --- | --- | --- | --- | --- |
| STRVCTVRE | Genome-wide features | - | hg38 | Random forest | Clinvar | Clinvar, gnomAD, Kronenberg et al. 2018 | Yes | Decipher | [5] |
| X-CNV (*) | Genome-wide features | - | hg19 | XGboost | dbVar | dbVar | No | ClinGen, Decipher, CNV sets mapping cancer predisposition genes | [6] |
| TADA (*) | Knowledge-based & genome-wide features | Haploinsufficiency (HI) index; gene list from the DDG2P | hg19 | Random forest | Decipher | Audano et al. 2019, gnomAD, UK Biobank, Aguirre et al. 2019, DGV | Yes | Clinvar | [7] |
| CADD-SV | Knowledge-based & genome-wide features | Gene list from the DDG2P | hg38 | Random forest | Simulated variants (proxy-pathogenic) | Human and chimpanzee-derived SVs (proxy-neutral) | Yes | Clinvar, ICGC, GTEx | [8] |
| ClassifyCNV | ACMG/ClinGen guidelines | ACMG/ClinGen guidelines | hg19 | - | - | - | - | - | [4] |

The explicit or implicit reliance of CNV assessment methods on the available sets of clinically resolved CNVs raises two major questions: (i) whether this may ultimately lead to a self-reinforced ascertainment bias towards genomic regions overlapping with the corpus of previously described disease-associated genes and pathogenic CNVs, and (ii), whether the existence of such a corpus nevertheless allows the identification of unbiased sequence and structural features of pathogenic and benign CNVs that can be generalized in a fair manner to the assessment of less well-represented CNV types in which WGS-based calling is currently enriched, such as short CNVs, CNVs involving genes not phenotypically relevant for the clinical case considered and CNVs involving only non-coding regions.

We addressed these questions, by first considering the characteristics of current reference databases of human CNVs in terms of the size and impact of these CNVs on protein-coding genes. As expected, we identified a systematic enrichment of pathogenic CNVs in longer variants affecting disease-associated genes. We then developed CNVscore, a supervised-learning approach for CNV scoring providing pathogenicity scores coupled with an estimate of the uncertainty on the prediction. The CNVscore framework combines tree ensembles and a Bayesian classifier trained on pathogenic and non-pathogenic CNVs from public databases. CNVscore was designed as a bundle of 23 independent classifiers, each trained in a leave-one-chromosome-out manner across autosomes and chromosome X, and on length-matched sets of pathogenic and non-pathogenic CNV sets, so as to prevent bias in predictions in later applications. The Bayesian component of CNVscore couples the pathogenicity predictions with a posterior probability that is transformed into an uncertainty score. The uncertainty and pathogenicity scores of CNVscore are orthogonal, making it possible to define subsets of CNVs for which the pathogenicity score can be considered to be of high, moderate or low reliability, reflecting the representativeness of the training set for the query variants.

We assessed the ability of CNVscore to distinguish between pathogenic and benign CNVs, by performing a comprehensive comparative benchmark test across independent CNV sets and against reference methods. We then investigated whether CNVs associated with CNVscores of low uncertainty were predicted significantly more accurately than those with high uncertainty scores. The performance of currently available scoring approaches was then assessed in challenging scenarios involving CNV sets with unconventional features, such as an absence of protein-coding elements in pathogenic CNVs or the presence of disease genes among benign variants. Finally, we evaluated the utility of the CNVscore framework for assessing the suitability of alternative training sets for scoring the French National Database of Constitutional CNVs, which contains annotated CNVs from clinical diagnosis.

## Results

### Characteristics of current reference databases of human CNVs in terms of the length of CNVs and their impact on protein-coding genes

We obtained a high-confidence non-redundant set of *n*=4368 and *n*=1565 pathogenic CNVs from the Clinvar and Decipher databases, respectively, and *n*=286,464 benign CNVs from reference sets (**Methods, Supplementary Table 1**). The distribution of genomic CNV lengths across the different sets largely reflected the resolution of the associated detection technologies (**Figure 1A** and **Supplementary Figure 1A**). Thus, pathogenic and likely pathogenic CNVs from the Decipher dataset were the largest variants (median 576,646 base pairs, bp), whereas the benign variants detected by short- and long-read sequencing were the shortest (median 218 bp, Wilcoxon one-tailed test *p*-value < 2.2e-16). However, the differences between pathogenic CNVs from Clinvar (median 6,717 bp) and benign variants from the Decipher Control and DGV sets (median 8,604 bp) were less striking, although still significant (*p* values of 5.79e-19 and 2.23e-3, respectively). We then classified CNVs according to the type of functional elements found within their genomic boundaries (**Figure 1B** and **Supplementary Figure 1B**). More than 92% of pathogenic CNVs and 79% of likely pathogenic CNVs, respectively, targeted Mendelian disease genes, with an additional ∼1-2% overlapping with enhancers of Mendelian disease genes mapping outside the CNV boundaries. The involvement of Mendelian disease genes also contrasted slightly with that for benign variants, in which pathogenic and likely pathogenic CNVs accounted for less than 16% and 4%, respectively, of the variants. Conversely, more than 45% of benign CNVs had no impact on Mendelian disease genes, either directly or via enhancers, and no impact on protein-coding exons, whereas this was the case for <2% of pathogenic and likely pathogenic variants. The fraction of CNVs with no obvious impact on protein-coding genes increased to 53-57% for datasets based exclusively on next-generation sequencing. Similar trends were observed when deletions and duplications were represented separately (**Supplementary Figure 1**). These findings indicate that the human pathogenic CNVs identified to date are strongly enriched in large variants affecting disease-associated genes.

**Figure 1.**
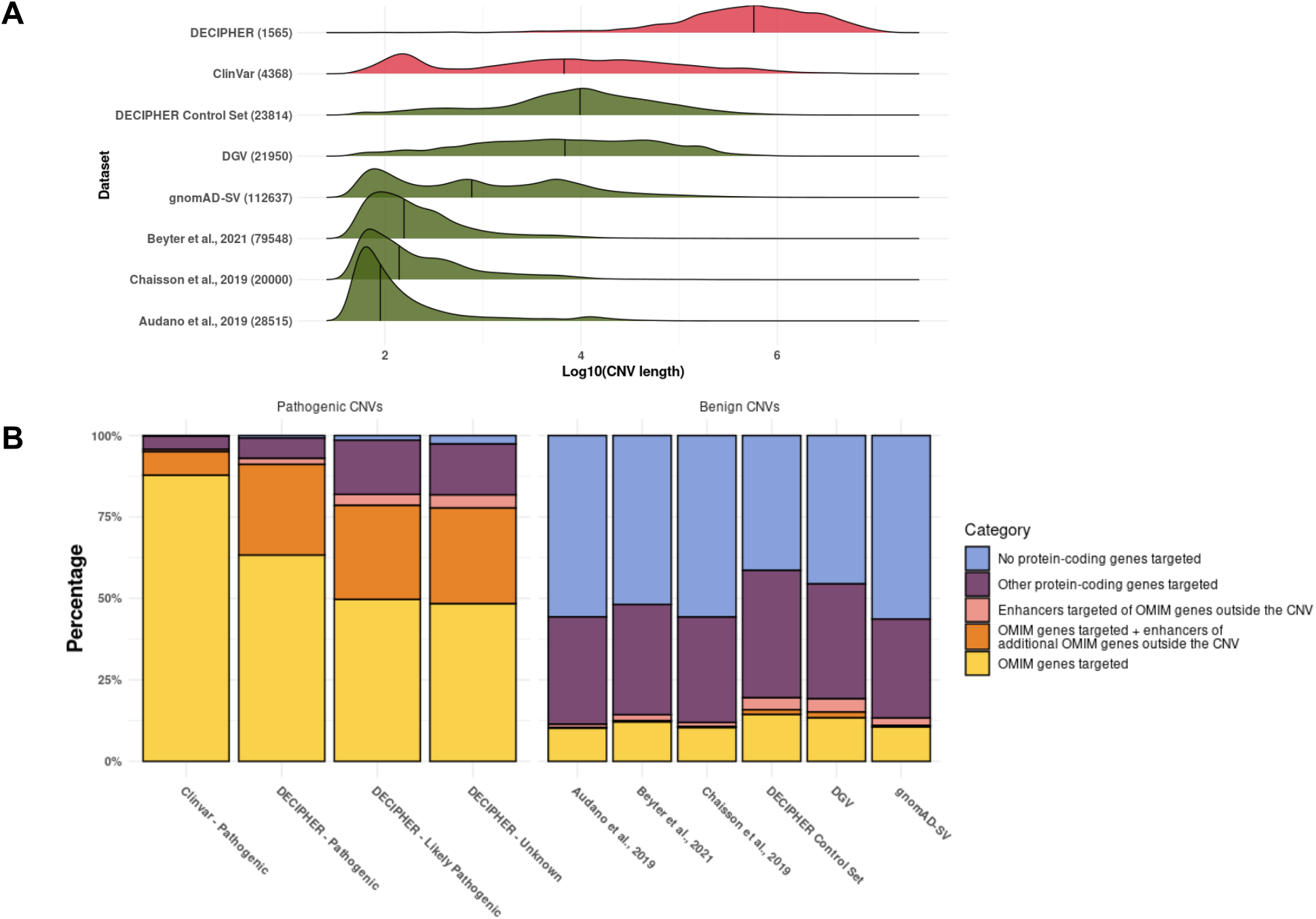
Genomic-length distribution and gene functional elements affected by reference pathogenic and benign CNV sets. (A) The CNV genomic length distribution (log-10 transformation of the number of base pairs) across pathogenic (red) and non-pathogenic (green) high-confidence non-redundant reference CNV sets (**Methods**). The median values are indicated with a vertical bar. The total number of CNVs within each set, aggregating deletions and duplications, is indicated in parentheses. (B) Bar plots showing the percentage of CNVs within each reference set belonging to five different categories, according to the type of functional elements targeted. Categories were considered mutually exclusive and included the following: (i) CNVs targeting Mendelian disease genes (referred to in the legend as “OMIM” genes for simplicity; yellow); (ii) CNVs targeting Mendelian disease genes and enhancers of other Mendelian disease genes mapping outside the CNV boundaries (orange); (iii) CNVs with no direct impact on Mendelian disease genes, but with an indirect impact through enhancers of Mendelian disease genes (pink); (iv) CNVs with no impact on Mendelian disease genes, either direct or through enhancers, but with an impact on other protein-coding exons (dark violet); (v) CNVs with no impact on Mendelian disease genes, either direct or through enhancers, and no impact on any other protein-coding exons (light violet). CNV sets were grouped according to their identification as pathogenic (left) or benign (right; **Methods**).

### Implementation of a supervised-learning strategy aiming to mitigate downstream ascertainment bias

The enrichment in large CNVs targeting Mendelian disease genes described above is challenging for supervised-learning approaches naively trained on such biased sets, in terms of (i) the identification of shorter pathogenic CNVs, (ii) avoiding an ascertainment bias driven by the partial genomic overlap between training CNVs and CNVs newly observed at the time of testing, and (iii) generalizing, with confidence, the identification of pathogenic CNVs characterized by combinations of features poorly represented in the training set, particularly as concerns protein-coding genes. We addressed these aspects as follows: We first built a training set in which the positive (i.e. pathogenic CNVs) and negative (i.e. benign CNVs) sets were matched for genomic length in a 1:1 ratio (**Methods**), as previously described (5–7). For constitution of the training set, we initially preferred Clinvar pathogenic CNVs over Decipher pathogenic CNVs, as the sample size was larger and the length distribution of these CNVs was closer to that for benign CNVs detected by whole-genome sequencing (**Figure 1A**). We, thus, matched *n*=3743 deletions and *n=*196 duplications from the Clinvar pathogenic CNV set with identical numbers from the benign CNV set, resulting in distributions matched for length (median of 4,499 bp for pathogenic deletions and 4,488 bp for benign deletions, and median of 17,338 bp for pathogenic duplications and 17,350 bp for benign duplications; **Supplementary Figure 2**). We then adopted a supervised-learning strategy characterized by a *leave-one-out* approach applied to chromosomes, as in Strvctvre [5], the only previous CNV scoring method to have adopted such a strategy, to our knowledge. Here, rather than training a single model, we trained a bundle of 23 independent models, with one chromosome left out in each (**Methods**). In this way, the downstream contamination of training and testing CNVs was prevented by design.

We minimized circularity and ascertainment bias, by deliberately ignoring CNV features based on prior clinical genetic evidence (e.g. genomic overlap with pathogenic CNVs, disease-associated genes or pathogenic SNVs), i.e. the evidence commonly used by medical guidelines for CNV evaluation [10] which has served as the basis of expert classifications. By contrast, CNVs were annotated with a total of *n*=38 genome-wide features not explicitly conveying associations of the targeted genomic elements with disease based on prior human knowledge (**Methods, Supplementary Table 2**). As expected, the distribution of these features across the pathogenic and non-pathogenic sets matched for length as described above showed that pathogenic CNVs were significantly enriched in variants targeting conserved genes, dosage-sensitive genes and genes intolerant to variants with a major functional impact, as demonstrated by various proxy variables (**Figure 2** and **Methods**). However, enrichment was more pronounced across deletions than across duplications, reflecting differences in pathogenic molecular mechanisms as well as sample size.

**Figure 2.**
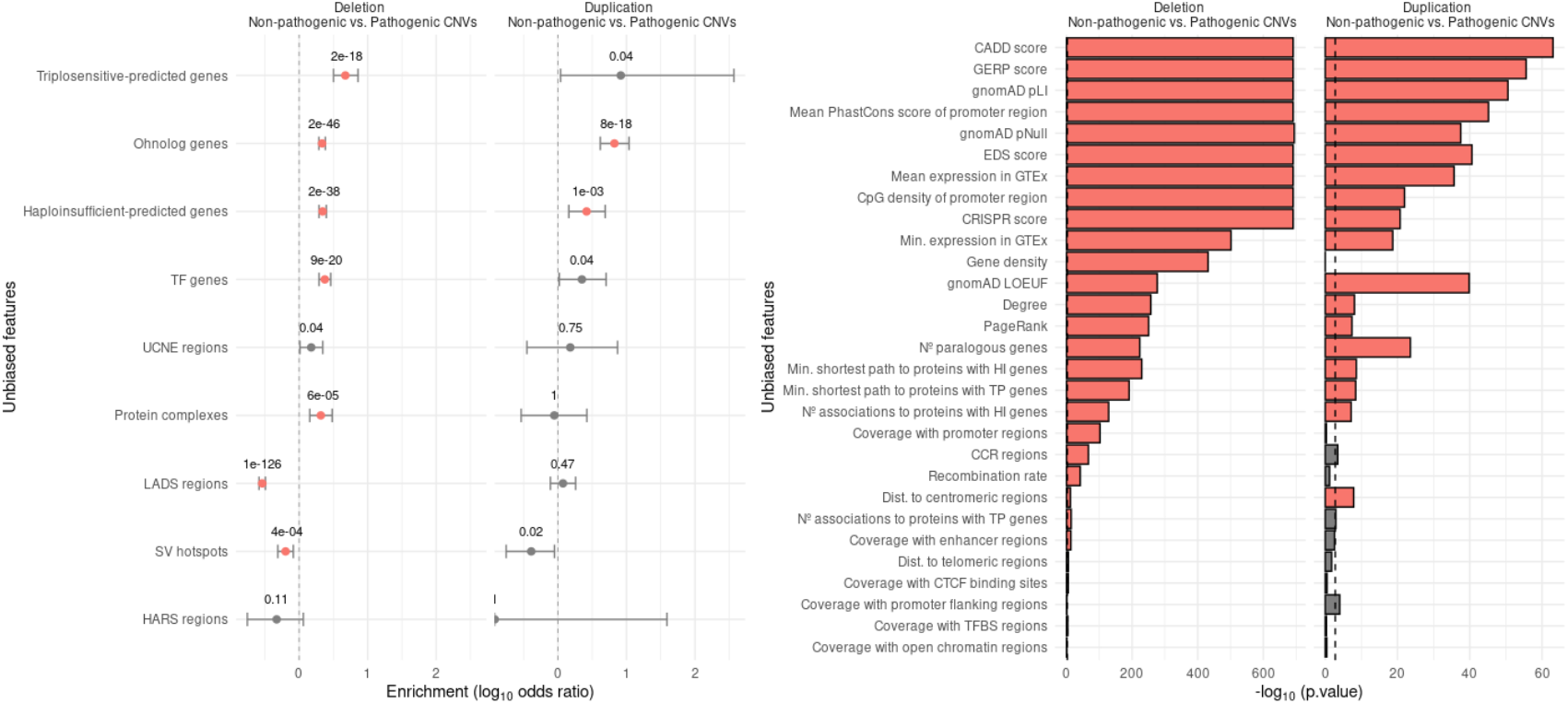
Relative distribution of the genome-wide features used by CNVscore between the pathogenic and benign CNV sets matched for length. Enrichment in categorical (A) and continuous (B) features was evaluated with Fisher’s exact tests and two-tailed Wilcoxon rank-sum tests, respectively. Within each panel, the results are shown separately for deletions (left side) and duplications (right side). The type I error rate was set to 0.05, and significance was assessed by Bonferroni correction according to the number of tests, i.e. *p-*value < 0.0028 (corresponding to 2 × 9 tests, for categorical features) *p*-value < 0.0022 (corresponding to 2 × 23 tests, for continuous features). Features with significant *p*-values are shown in orange. Point estimates and 95% confidence intervals for the log_10_ odds ratios are shown in (A), in which positive values indicate enrichment in a particular feature for pathogenic CNVs relative to benign CNVs. Minus log_10_ Wilcoxon *p*-values are represented in (B) as bar plots. Feature names and acronyms are described in **Supplementary Table 2**.

### Development of CNVscore, a pathogenicity scoring system coupled with uncertainty estimates

The enrichment of pathogenic CNVs in known disease-associated genes and specific gene-based features, raised the question as to whether it was feasible to generalize a supervised-learning approach to the assessment of CNV types poorly represented in the training set, such as: (i) pathogenic CNVs encompassing genes not previously known to be associated with disease, (ii) CNVs that are non-pathogenic despite including disease-associated genes, and (iii) pathogenic CNVs containing only non-coding regions. We reasoned that a Bayesian approach could provide not only a pathogenicity score, but also a quantitative assessment of the uncertainty or reliability of the pathogenicity scoring of a CNV, based on how often the features of the CNV are observed across the training set. To this end, we developed CNVscore, a supervised-learning approach combining tree ensembles and Bayesian classifiers (**Figure 3, Methods**). Two independent CNVscore models were trained: one for deletions and the other for duplications. For each model, we first trained a gradient-boosting model on the training set described above, using the *n*=38 previously reported features. Gradient boosting fits a set of decision trees [11], each implementing a concatenation of decision rules for a subset of quantitative and qualitative features. Throughout the training process, the order of features evaluated and their decision thresholds are established so as to optimize predictive performance on the training set. A majority voting-like strategy gathers the output of each tree, to generate a final prediction. However, unlike random forest approaches, the trees in gradient boosting are not independently trained, but trained in a successive manner. Thus, each additional tree optimizes the predictive performance for those instances of the training set for which the previous trees tended to fail.

**Figure 3.**
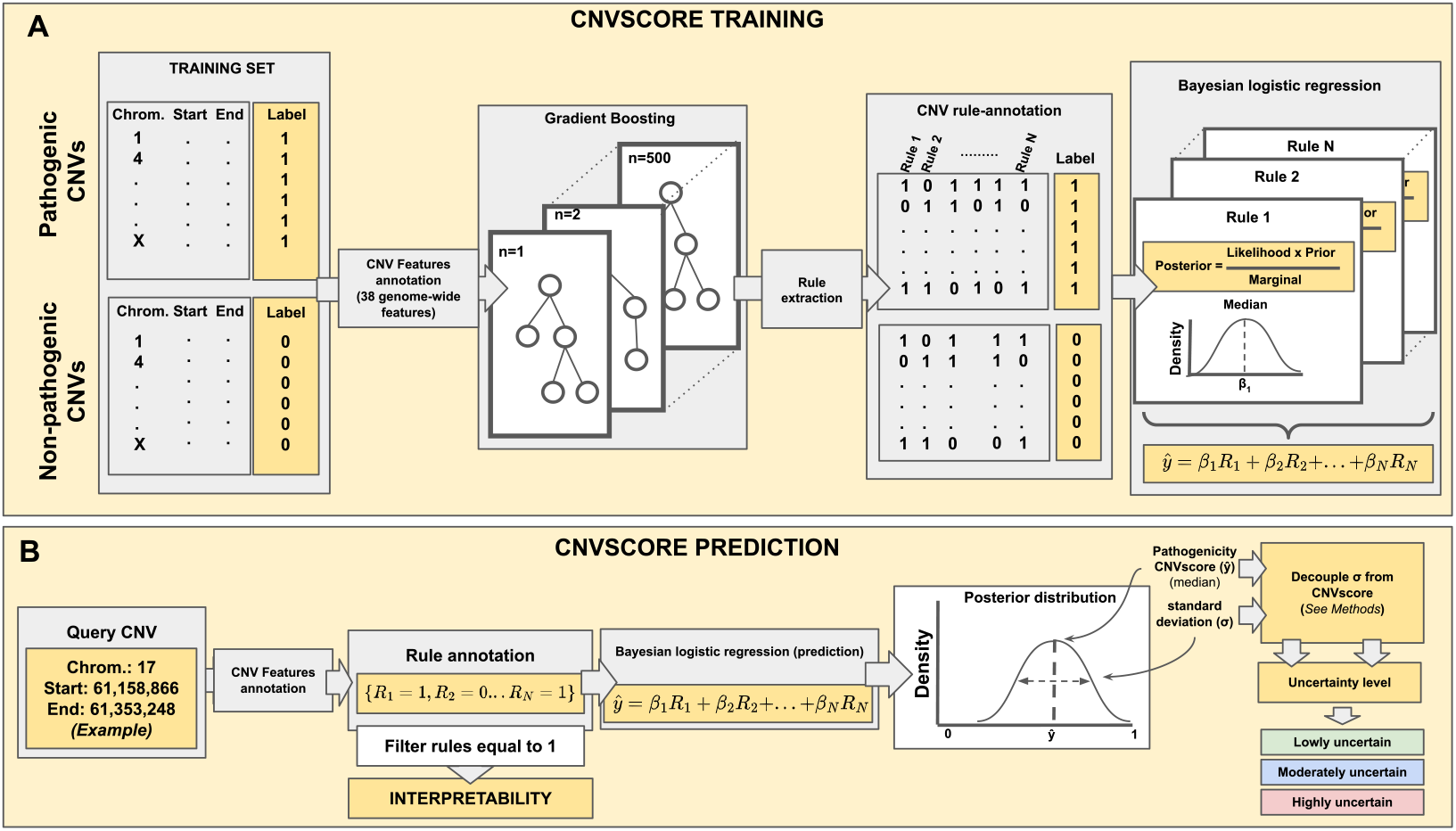
Schematic representation of the CNVscore method. (A) CNVscore training phase: A gradient-boosting model was first trained to classify pathogenic and benign CNVs on 38 genome-wide features. Each of the resulting trees was decoupled into a set of independent decision rules, which were used to annotate CNVs in a binary manner. Such vectors were used as input, to train a Bayesian generalized linear regression model on the same CNV sets. The likelihoods of the model parameters were combined with priors to generate their posterior probability. (B) Scoring of query CNVs: Once trained, the Bayesian model calculates, for each query CNV, a posterior probability distribution indicating the probability of the CNV being pathogenic. The centrality measurement of the posterior distribution of pathogenicity is considered as the *pathogenicity* CNVscore, each such score being associated with a quantitative measure of the degree of confidence (i.e. dispersion) of the prediction, considered as the *uncertainty* CNVscore. Two independent CNVscore models were trained for deletions and duplications, respectively (**Methods**).

After the first training phase, each of the resulting trees was decoupled in a set of independent decision rules, which were then used to annotate the CNVs in the training set in a binary manner (“1” or “0”), depending on whether or not they followed each rule. Each CNV in the training set was, therefore, associated with a binary vector of *n* dimensions, where *n* is the number of rules derived from the trees in the gradient-boosting model. These vectors were then used as feature vectors, to train a Bayesian generalized linear regression model on the same CNV sets (**Figure 3A**). Under a Bayesian framework, the coefficients of the regression are set as a probability distribution rather than as a fixed-value estimate. The likelihoods of the model parameters (i.e. the probability of observing the data given that the underlying parameter values are true) were combined with their prior probabilities by Bayes’ theorem, to generate posterior probability distributions (i.e. the probability that the parameters are equal to a given value after observation of the data; **Methods**). The trained Bayesian model then calculated, for each query CNV, a posterior probability distribution indicating the probability of the CNV being pathogenic (**Figure 3B**). The posterior distributions for each CNV were then summarized, using the median as a measurement of centrality, and the standard deviation (sd) as a measurement of dispersion (**Methods)**. Thus, for each CNV evaluated, the centrality of the posterior distribution of pathogenicity was assessed by the *pathogenicity* CNVscore, with each such score being associated with a quantitative measurement of the degree of confidence (i.e. dispersion) of the prediction, which we call the *uncertainty* CNVscore. By design, the *pathogenicity* and *uncertainty* CNVscores are orthogonal, and therefore convey independent information (**Supplementary Figure 3; Methods**).

### Pathogenicity CNVscores had a good classification performance across independent sets of pathogenic and benign CNVs matched for length

Ten-fold cross-validation of CNVscore on the previously described training set of Clinvar pathogenic CNVs and benign CNVs (**Methods**) gave an AUROC of .945 and .885,and an AUPR of .913 and .868, for *n*=7,486 deletions and *n*=392 duplications, respectively (**Table 2**). These figures indicate a better performance for deletions than for duplications, reflecting their training set size and more challenging assessment. However, they also show that a Bayesian model coupled with a gradient-boosting model outperforms the gradient-boosting model alone (**Supplementary Figure 4**). CNVscore reached values similar to those obtained with alternative supervised-learning methods trained on analogous sets of pathogenic and benign CNVs (Strvcture, X-CNV and TADA). This consistency can be partly explained by the strong correlations between the results of these methods (Spearman-rank r = 0.781, 0.744 and : 0.742, for Strvcture, X-CNV and TADA, respectively, with significant *p*-values< 2.2e-16 across the 3 comparisons). Conversely, supervised-learning methods based on simulated CNVs (e.g. CADD-SV), and medical guidelines (e.g. ClassifyCNV) had the poorest performances (**Table 2**).

**Table 2.**
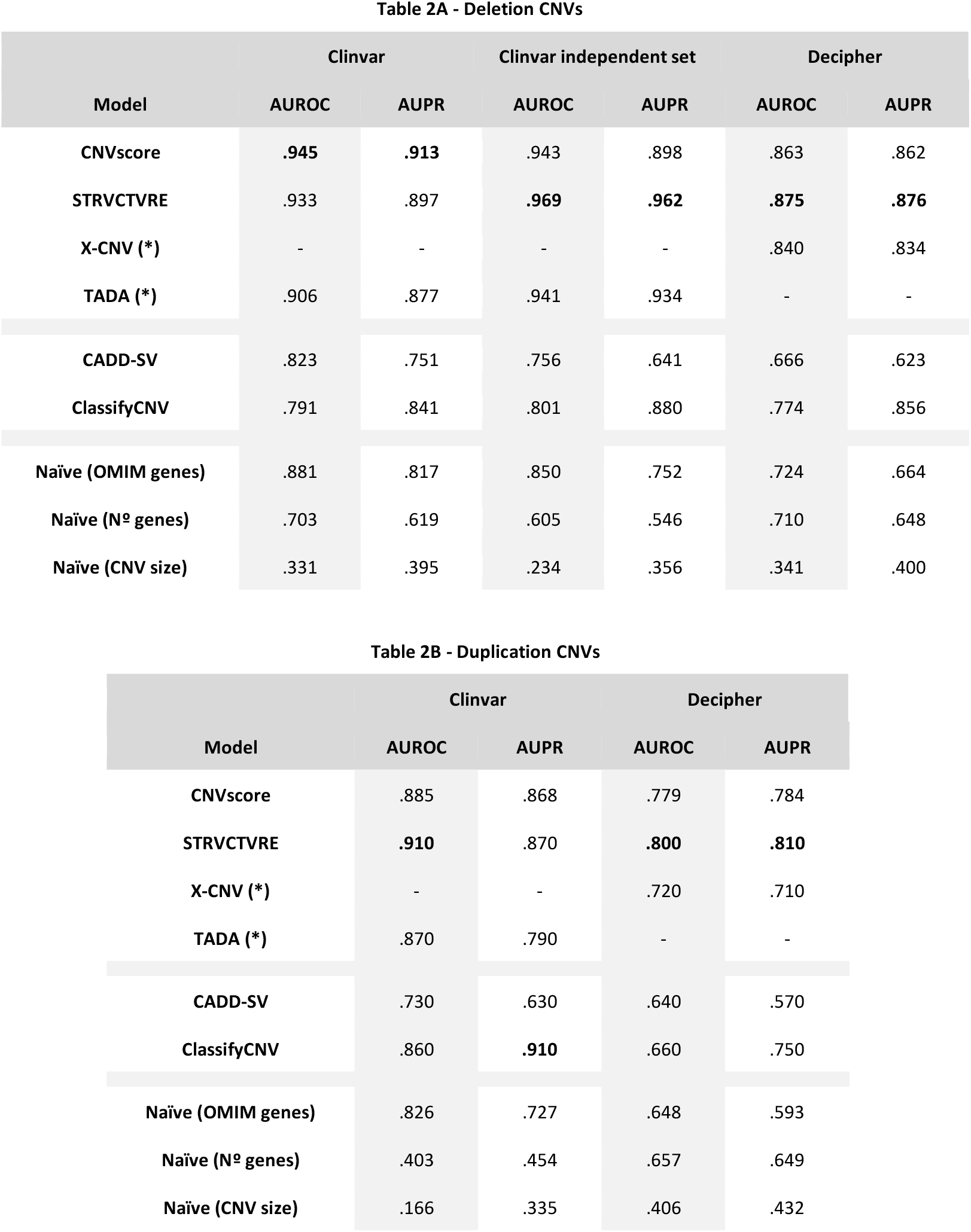
Comparative benchmarking of the classification into pathogenic and benign CNVs across different methods and datasets. Tables represent the area under the receiver operating characteristics curve (AUROC) and area under the precision-recall curve (AUPR) values obtained by CNVscore together with five alternative methods and three naïve control approaches across deletions (A) and duplications (B) (**Methods**). Three sets were considered, corresponding to Clinvar, Clinvar independent (>2021), and Decipher (see text and **Methods** for details). The Clinvar independent set was not assessed for duplications because the sample size was too small (**Supplementary Table 3**). Values in bold indicate the highest AUROC and AUPR values across methods within a dataset. Asterisks (*) indicate methods (TADA and X-CNV) with potential contamination between their training sets and the benchmark CNV sets evaluated here (see **Table 1**). Dashes (-) indicate values masked because of such contamination. The full data are reported in **Supplementary Tables 4, 5, 6, 7 and 8**.

Nevertheless, when interpreting these findings, it is important to bear in mind the observed enrichment of pathogenic CNVs in variants targeting Mendelian disease genes and, more generally, protein-coding genes. We, therefore, independently calculated the classification performance that would be achieved by these two simple features, as a baseline. Here, the presence/absence of Mendelian disease genes within the CNV would give an AUROC of .881 and .826, and an AUPR of .816 and .727, for deletions and duplications, respectively. The number of protein-coding genes within the CNV would give an AUROC of .702 and .403, and an AUPR of .618 and .454, for deletions and duplications, respectively. The trends identified were consistently reproduced in three independent tests: one based on Clinvar pathogenic deletions reported since 2021, and the other two on sets of Decipher pathogenic CNVs (deletions and duplications, respectively; **Table 2**). Our findings indicate that pathogenic CNVscore has a good classification performance, comparable to that of alternative supervised-learning methods, and generalizable to independent CNV datasets. However, its performance constitutes an improvement in accuracy of only ∼7% relative to that achieved with a single feature, such as Mendelian disease gene content.

### Low-uncertainty CNVscores identify subsets of CNVs associated with a higher pathogenicity classification accuracy

The Bayesian framework implemented by CNVscore made it possible to associate an uncertainty score to each pathogenicity score (**Methods**). These two scores provide independent information (Spearman’s rank correlation = -0.0042 and -0.012, *p*.value = 0.714 and 0.809, for deletions and duplications, respectively). We therefore further split the previously matched-by-length CNV sets according to their *uncertainty* CNVscores, into low-, moderate- and high-uncertainty classes (**Methods**). Higher AUROC values were obtained for low-uncertainty than for high-uncertainty CNVs: .948 vs .937, and .942 vs .767, for the Clinvar and Decipher deletion sets, respectively, with significant *p-*values <1e-33 in both cases (**Methods**). Similar trends were observed for duplications: .994 vs .759, and .856 vs .572, for analogous comparisons, respectively, with significant *p*-values <2.56e-34 in both cases (**Table 3, Methods**). The association of low *uncertainty* CNVscores with high classification performance was also generalized to alternative CNV supervised-learning scores, such Strvcture, TADA, X-CNV and CADD-SV. All four methods performed significantly better with low-uncertainty CNVs, across both the Clinvar and Decipher sets, and for both deletions and duplications, in all cases other than the use of CADD-SV on Decipher deletions (**Table 3**). The associations were observed despite the lack of correlation between *uncertainty* CNVscore and the results of these alternative methods (Spearman’s rank correlation coefficients of -0.059 to 0.065 for all comparisons evaluated). It was not possible to carry out similar tests on Clinvar deletions reported since 2021, due to the small size of the sample (**Supplementary Table 3**). These results demonstrate the capacity of a Bayesian uncertainty score to identify subsets of CNVs for which pathogenicity CNVscores and alternative supervised-learning methods yielded a higher classification accuracy.

**Table 3.**
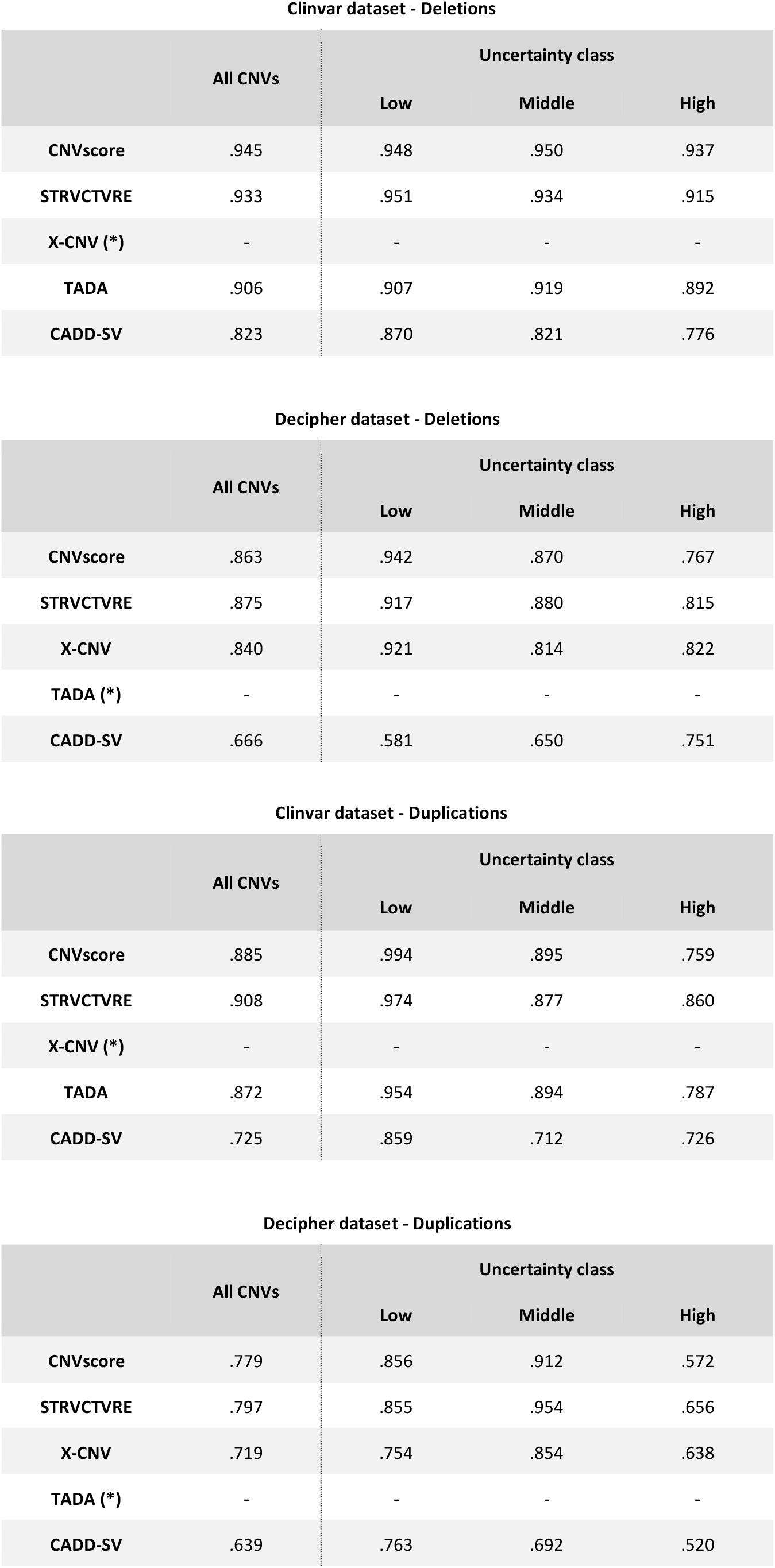
Classification performance across low-, medium- and high-uncertainty CNV subsets. The tables represent the area under the receiver operating characteristics curve (AUROC) obtained with CNVscore, together with four alternative supervised-learning methods across deletions and duplications (**Methods**). Two sets were considered, corresponding to the Clinvar and Decipher sets (see text and **Methods** for details). AUROC values are shown for the total CNV sets (indicated as “All”), and for the corresponding CNV subsets split on the basis of *uncertainty* CNVscores, into low-, medium- and high-uncertainty classes (**Methods**). Asterisks (*) indicate methods (TADA and X-CNV) with potential contamination between their training sets and the benchmark CNV sets evaluated here (see **Table 1**). Dashes (-)indicates values masked because of such contamination. The full data are reported in **Supplementary Tables 9, 10, 11 and 12**. The number of CNVs within each uncertainty level for Clinvar and Decipher, for deletions and duplications, is reported in **Supplementary Table 13**. Analogous tests could not be carried out for Clinvar deletions reported since 2021, due to the small sample size.

**Clinvar dataset - Deletions**
|  | All CNVs | Uncertainty class |  |  |
| --- | --- | --- | --- | --- |
|  |  | Low | Middle | High |
| CNVscore | .945 | .948 | .950 | .937 |
| STRVCTVRE | .933 | .951 | .934 | .915 |
| X-CNV (*) | - | - | - | - |
| TADA | .906 | .907 | .919 | .892 |
| CADD-SV | .823 | .870 | .821 | .776 |

**Decipher dataset - Deletions**
|  | All CNVs | Uncertainty class |  |  |
| --- | --- | --- | --- | --- |
|  |  | Low | Middle | High |
| CNVscore | .863 | .942 | .870 | .767 |
| STRVCTVRE | .875 | .917 | .880 | .815 |
| X-CNV | .840 | .921 | .814 | .822 |
| TADA (*) | - | - | - | - |
| CADD-SV | .666 | .581 | .650 | .751 |

**Clinvar dataset - Duplications**
|  | All CNVs | Uncertainty class |  |  |
| --- | --- | --- | --- | --- |
|  |  | Low | Middle | High |
| CNVscore | .885 | .994 | .895 | .759 |
| STRVCTVRE | .908 | .974 | .877 | .860 |
| X-CNV (*) | - | - | - | - |
| TADA | .872 | .954 | .894 | .787 |
| CADD-SV | .725 | .859 | .712 | .726 |

**Table 3. Classification performance across low-, medium- and high-uncertainty CNV subsets.** The tables represent the area under the receiver operating characteristics curve (AUROC) obtained with CNVscore, together with four
|  | All CNVs | Uncertainty class |  |  |
| --- | --- | --- | --- | --- |
|  |  | Low | Middle | High |
| CNVscore | .779 | .856 | .912 | .572 |
| STRVCTVRE | .797 | .855 | .954 | .656 |
| X-CNV | .719 | .754 | .854 | .638 |
| TADA (*) | - | - | - | - |
| CADD-SV | .639 | .763 | .692 | .520 |

### Classification of performance in challenging scenarios

As described above, CNVscore was trained on a set of pathogenic and benign CNVs matched for length. A leave-one-chromosome-out approach was used for training, which was based on genome-wide features not explicitly conveying knowledge-based information about the associations with disease of the targeted genes. Despite the preventive measures taken to mitigate ascertainment bias, the pathogenic CNVs in the training sets were enriched in CNVs targeting Mendelian disease genes and, more generally, protein-coding genes, relative to benign CNVs (**Figure 1B**). We show above that either of these two naïve features can itself be used to achieve a classification performance close to those of the supervised-learning scores across the different sets evaluated (**Table 2**). We, thus, characterized the behavior of these features while controlling for such biases in the CNV evaluation sets. We investigated four different evaluation scenarios by sampling deletions from the pathogenic set and the benign CNV set fulfilling specific requirements, as follows: (i) Scenario #1, in which only pathogenic and benign CNVs targeting Mendelian disease genes were considered; (ii) Scenario #2, in which only pathogenic and benign CNVs not targeting Mendelian disease genes were considered; (iii) Scenario #3, in which pathogenic CNVs not targeting Mendelian disease genes and benign CNVs targeting Mendelian disease genes were considered, and (iv) Scenario #4, in which pathogenic and benign CNVs not targeting protein-coding genes were considered. For each scenario, two sets were obtained, with either the Clinvar or the Decipher pathogenic CNV set used as the initial set (**Methods**).

Supervised-learning methods trained on clinically annotated samples, (CNVscore, Strvcture and X-CNV) were robust overall if the presence/absence of Mendelian disease genes across the testing sets was controlled for (**Table 4**, Scenarios #1 and #2), but with AUROC and AUPR values slightly lower than those for the biased sets. Significant decreases in performance were, nevertheless, observed for two methods: CADD-SV, a method trained on simulated variants as a proxy for pathogenic variants, and ClassifyCNV, a method based on medical guidelines, which behaved as close-to-random classifiers in Scenarios #1 and #2, respectively (**Table 4**). However, under the more challenging setting represented by Scenario #3, all the methods evaluated had a significantly lower capacity to distinguish pathogenic CNVs not targeting Mendelian disease genes from benign CNVs targeting Mendelian disease genes, even behaving as random classifiers in the case of the Decipher set. Similarly, extremely poor performances were obtained for Scenario #4, for which none of the methods, including CNVscore, was able to classify CNVs not targeting protein-coding genes. Consistent with the results reported above, the classification performances observed across the challenging scenarios were, overall, inversely correlated with the fraction of CNVs annotated with a *high-uncertainty* CNVscore (**Figure 4**). Thus, Decipher-based scenarios, which had a higher percentage of CNVs within the high-uncertainty class than those based on Clinvar, resulted in the lowest AUROC values, for all the methods evaluated.

**Table 4.**
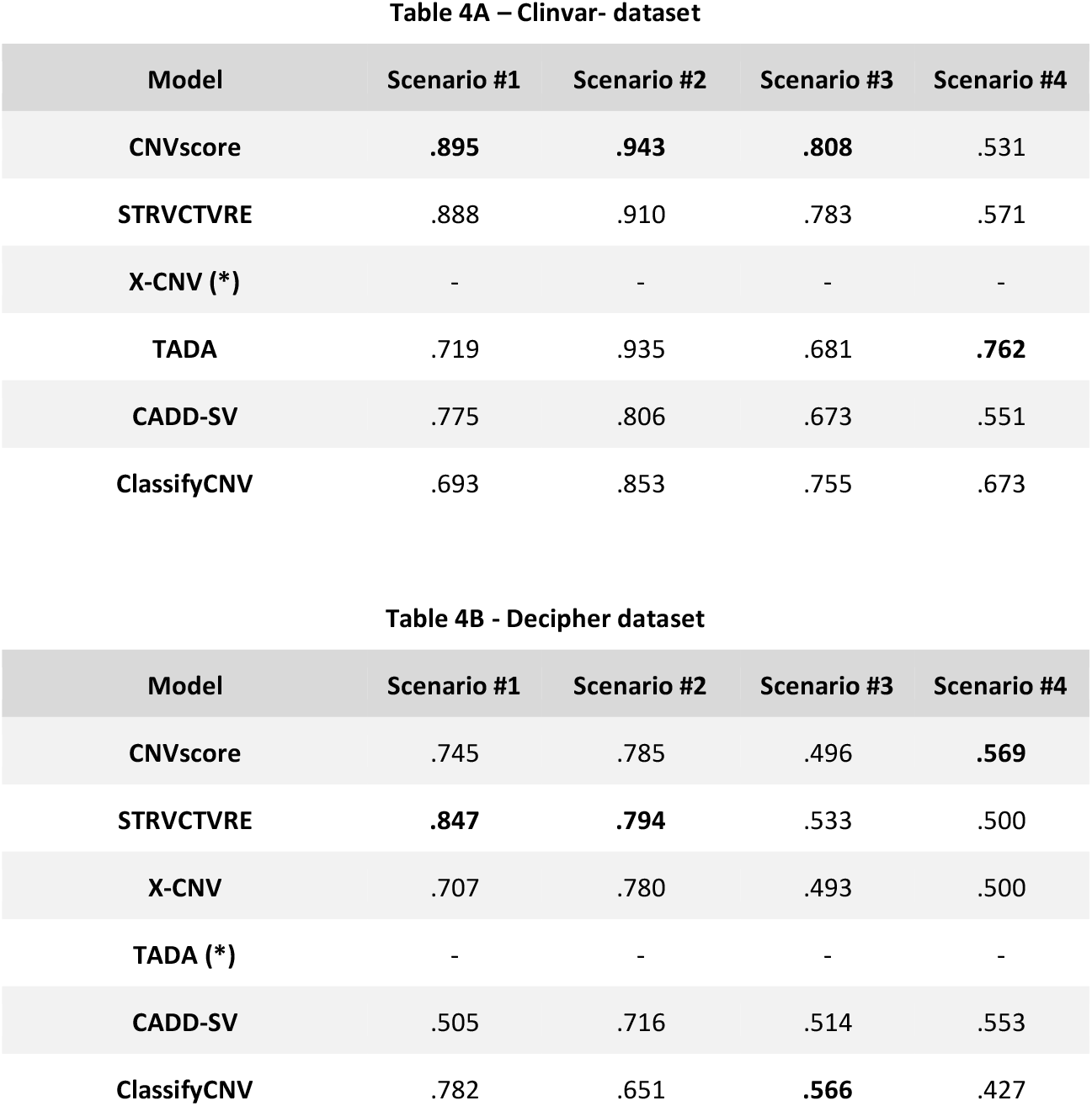
Classification performance in challenging scenarios. The tables represent the area under the receiver operating characteristics curve (AUROC) obtained with CNVscore, together with five alternative methods across deletions (**Methods**). Two reference sets were considered, corresponding to the Clinvar and Decipher sets, subsets from which were sampled to mimic four different scenarios (see text and **Methods** for details). Asterisks (*) indicate methods (TADA and X-CNV) with potential contamination between their training sets and the benchmark CNV sets evaluated here (see **Table 1**). Dashes (-) indicate values masked because of such contamination. The full data are reported in **Supplementary Tables 14 and 15**. Analogous tests could not be carried out on duplication sets because of the small sample size.

**Figure 4.**
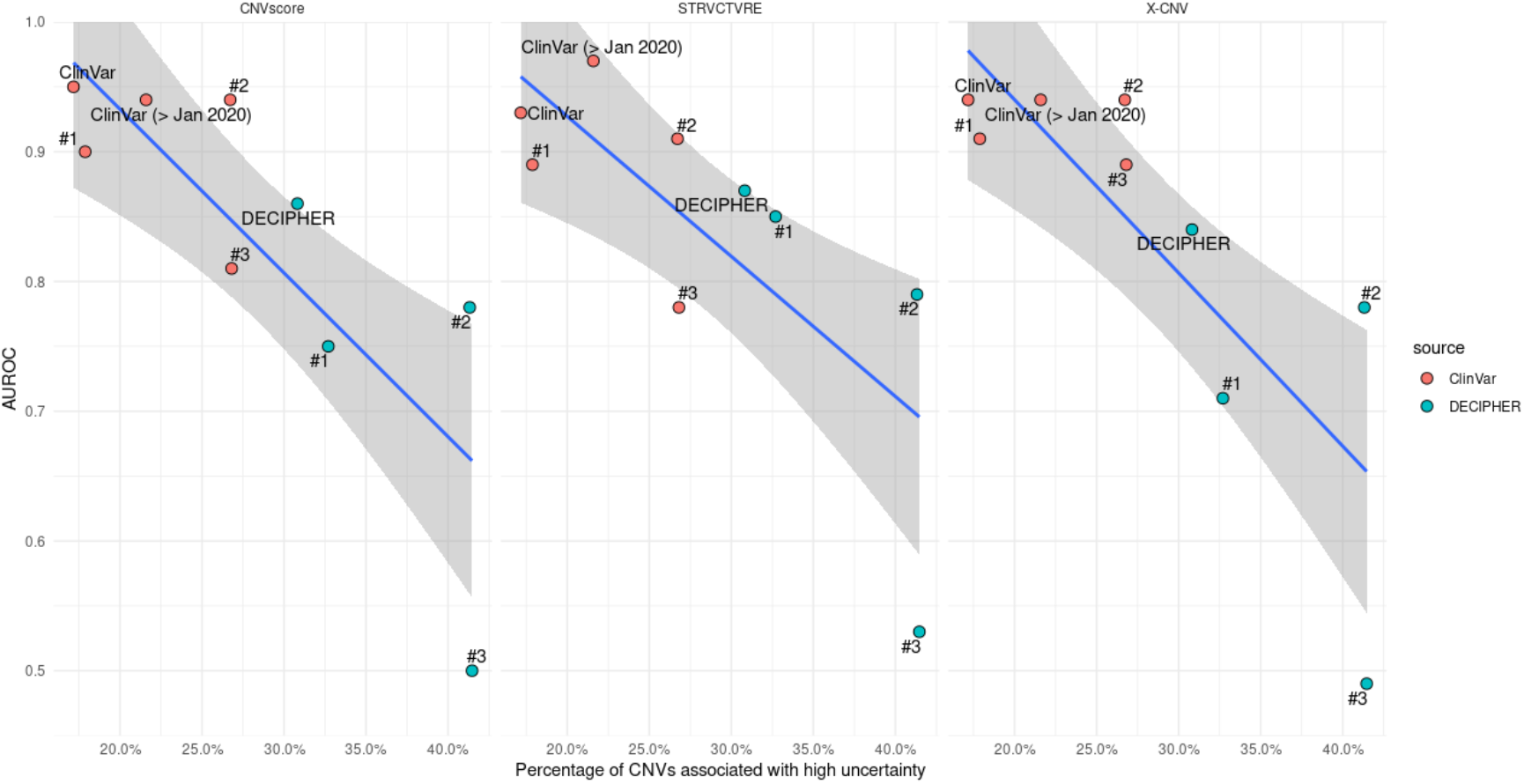
Classification performance across diverse evaluation sets as a function of the fraction of highly uncertain CNVs. This figure shows the classification performances (AUROC values, *y*-axis), of the three principal supervised-learning methods evaluated, i.e. CNVscore (left), Strvture (middle) and X-CNV (right), across different evaluation scenarios: #1, in which only pathogenic and benign CNVs targeting Mendelian disease genes were considered; #2, in which only pathogenic and benign CNVs not targeting Mendelian disease genes were considered; and #3, in which pathogenic CNVs not targeting Mendelian disease genes and benign CNVs targeting Mendelian disease genes were considered. For each scenario, two sets were obtained, using the Clinvar (red dots) or the Decipher (green dots) CNV set as the initial set (**Methods**). The *x*-axis shows the fraction of highly uncertain CNVs within each set, as calculated by CNVscore (**Methods**). Blue lines and shadowed areas represent the linear regression and 95% confidence intervals. Of note, scenario #4, described in the text, in which pathogenic and benign CNVs not targeting protein-coding genes were considered is not shown in this figure, because of the small sample size in both Clinvar (*n* = 14) and Decipher (*n* = 30; **Supplementary Table 3**). AUROC performances for these two sets are reported in **Table 4**.

**Figure 5.**
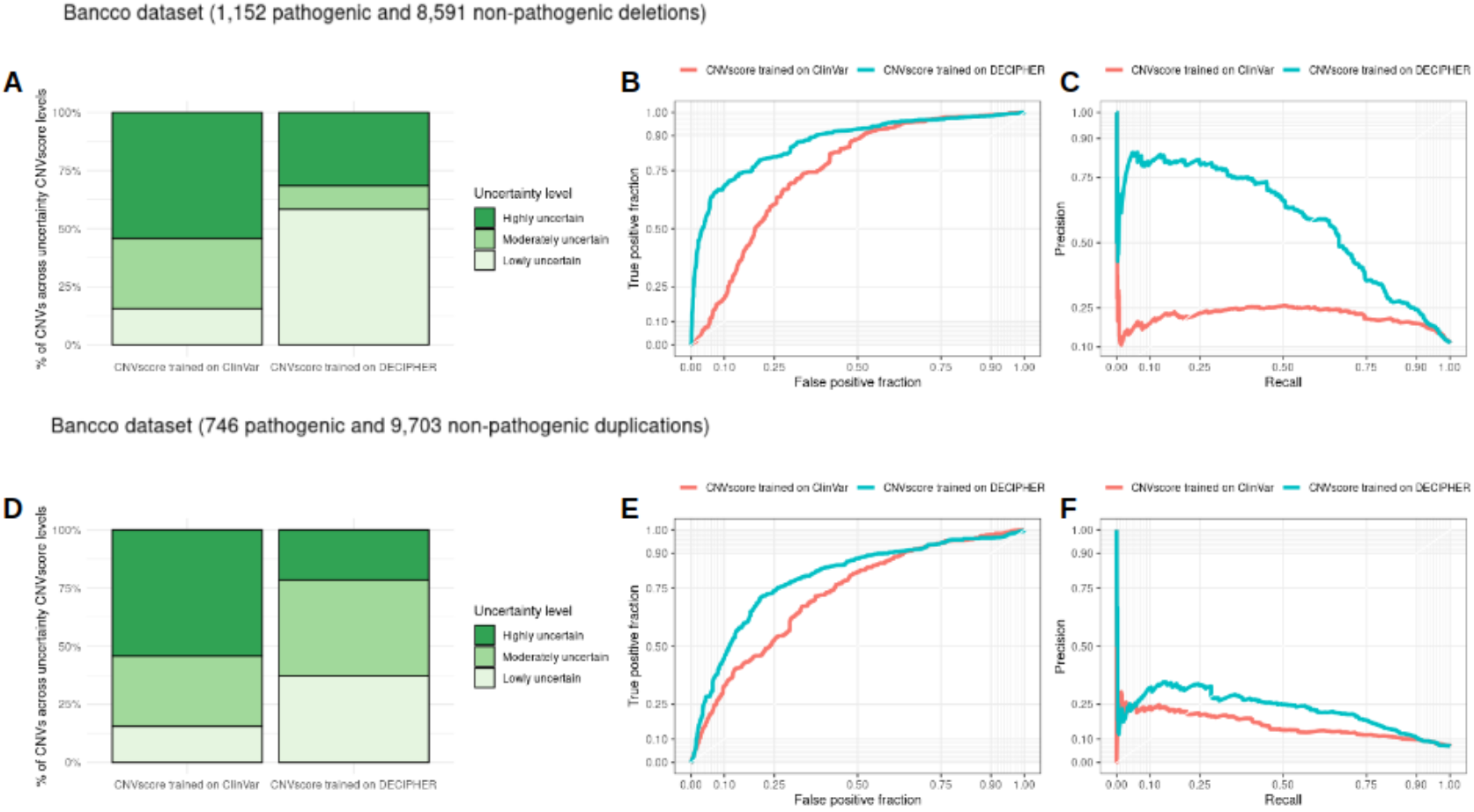
Application of the CNVscore framework to the French National Databank of Constitutional CNVs (BANCCO). The figure shows the results of the CNVscore framework on n=9,743 deletions (top panels) and *n*=10,449 duplications (bottom panels) identified through aCGH, mostly in the context of developmental delay diagnosis (http://bancco.fr). The left panels (**A, D**) show the fraction of CNVs assigned with low, moderate or high uncertainty by a CNVscore model trained on the Clinvar (left bar) or Decipher (right bar) datasets. The middle and right panels show the receiver operating characteristics curve (ROC) and the precision-recall curve (PR) obtained across deletions (top) and duplications (bottom), with a CNVscore model trained on the Clinvar (red curves) or Decipher (blue curves) datasets. For deletions (**B, C**), CNVscore trained on Decipher yielded an AUROC of .872 and an AUPR of .585; CNVscore trained on Clinvar yielded an AUROC of .746 and an AUPR of .219. For duplications (**E, F**), CNVscore trained on Decipher yielded an AUROC of .792 and an AUPR of .229; CNVscore trained on Clinvar yielded an AUROC of 0.720 and an AUPR of 0.160.

### Clinical utility of CNVscore in the French National Database of Constitutional CNVs

Finally, we evaluated the utility of the CNVscore framework on BANCCO, the French National Database of Constitutional CNVs. BANCCO gathers annotations from clinical diagnosis from 10 French university hospitals and two health consortia (the university hospitals of the Rhone-Alpes-Auvergne and of West regions, respectively). Here, we considered a total of *n* = 9,743 deletions and *n* = 10,449 duplications identified through aCGH, mostly in the context of developmental delay diagnosis (http://bancco.fr). The application of a CNVscore model trained on the Clinvar set revealed that a surprisingly high fraction of CNVs (54% for deletions and 49% for duplications) were associated with high-uncertainty CNVscores (**Figure 4**). Conversely, when a CNVscore model trained on the Decipher set was applied, a lower fraction of CNVs were assigned to the high uncertainty class (31% and 22%, for deletions and duplications, respectively). Consistent with the observed uncertainty levels, the pathogenicity CNVscores produced by a model trained on the Decipher dataset (**Methods**) gave a higher prediction accuracy (AUROC=.87 and .79; and AUPR=.58 and .23 for deletions and duplications, respectively) than those obtained with a model trained on the Clinvar dataset (AUROC=.75 and .72; and AUPR=.22 and .16 for deletions and duplications, respectively). These results highlight the capacity of CNVscore to provide an objective assessment of the suitability of alternative training sets for scoring a target clinical CNV dataset. Here, consistently with the focus of Decipher on the diagnosis of rare diseases, it appeared to be more representative as a training set for the BANCCO CNVs than Clinvar. Importantly, such assessments can be performed without prior knowledge of the actual clinical classification of the target CNVs, making it possible to perform model selection before the CNV scoring of clinically uncharacterized CNV sets identified in rare disease cohorts.

## Discussion

We used CNVscore, a supervised learning model combining gradient boosting with Bayesian logistic regression, for the classification of pathogenic and benign CNVs. Unlike alternative supervised-learning approaches, CNVscore combines a pathogenicity score with an estimate of uncertainty, making it possible to evaluate the suitability of the training set for the query variants. Pathogenicity CNVscores reached classification performances similar to those of state-of-the-art techniques in comparative benchmark tests across independent sets, in which duplications generally appeared to be more challenging to classify than deletion CNVs. Furthermore, CNVscore identified low-uncertainty CNV subsets for which supervised-learning approaches resulted in a higher classification accuracy. Finally, we identified challenging scenarios with enrichment in pathogenic CNVs not targeting protein-coding elements or benign variants encompassing disease-associated genes, for which current methods for CNV scoring, including *pathogenicity* CNVscore, presented major limitations.

Our findings show that supervised-learning methods are robust for distinguishing between pathogenic and benign CNVs involving coding regions, consistent with previous benchmarks [5–7], despite the biases in their associated training sets in terms of length and Mendelian disease gene content. However, two methods were found to be compromised: CADD-SV, a method trained on simulated variants as a proxy for pathogenic variants, which failed in a scenario in which only pathogenic and benign CNVs targeting Mendelian disease genes were considered; and ClassifyCNV, a method based on medical guidelines, which failed in a scenario in which only pathogenic and benign CNVs not targeting Mendelian disease genes were considered. However, all the methods struggled to prioritize atypical cases in comparisons of benign and pathogenic CNVs that did and did not target Mendelian disease genes, respectively. Furthermore, all the methods evaluated behaved as random classifiers when faced with CNVs not encompassing coding elements. These results raise concerns about the generalizability of current supervised-learning approaches for evaluating CNV types poorly represented in current reference Mendelian disorder datasets.

These observations highlight three major areas for further improving the classification accuracy of supervised-learning approaches. First, there is a need for more informative genome-wide features for distinguishing between pathogenic and non-pathogenic duplications; large-scale genome and transcriptome projects characterizing genomic regions that are tolerant to higher dosage would be useful in this respect [12]. Second, it is important to assess the consistency of the patient’s phenotype with the targeted genomic elements, as this can help prevent the misclassification as pathogenic of CNVs affecting Mendelian genes not associated with the clinical signs investigated. Indeed, this aspect has already been incorporated into current computational frameworks to assist the manual assessment of CNVs [3,13]; it would require the systematic incorporation of phenotypes into reference databases, such as Clinvar. Third, there is also a need for large-scale mutational scanning projects on cellular systems [14], to provide experimental evidence for CNVs not affecting coding genes. This aspect is especially relevant in light of the current scarcity of clinically annotated variants targeting only regulatory regions, which constitutes a major limitation for the development of supervised-learning approaches, as previously observed for non-coding SNVs [15].

The *pathogenicity* CNVscore method developed here can be used to prioritize large candidate sets of CNVs detected by WGS, to mitigate ascertainment bias towards large CNVs overlapping previously reported disease-associated genomic elements. However, we showed that classification accuracy across different evaluation settings was inversely correlated with the fraction of CNVs associated with *high*-*uncertainty* CNVscores. These results provide evidence that *uncertainty* CNVscores can be used for the analytical identification of subsets of CNVs with characteristics diverging from those of the training sets used in supervised-learning methods, leading to a drop in performance. Thus, *uncertainty* CNVscores may be used to filter candidate CNVs not well suited for assessment by supervised-learning methods trained on the currently available sets of clinically annotated CNVs. Furthermore, the fraction of CNVs associated with highly unreliable CNVscores may be used as a model-decision criterion, for identification of the most suitable training set for the scoring of CNV sets identified in genetically uncharacterized clinical cohorts. To facilitate the use of CNVscore, we have implemented an application programming interface (API) supporting remote queries. In addition, CNVscore is integrated into the CNVxplorer framework for the clinical assessment of CNVs identified in patients with rare diseases (http://cnvxplorer.com, [3]). The complete list of CNVs identified in a patient’s genome can, therefore, first be filtered by combining the pathogenicity and uncertainty CNVscores, to obtain a shortlist of high-confidence candidate pathogenic CNVs. This shortlist can then be further evaluated through in-depth manual assessment of the genomic, epigenomic and clinical annotations with tools such as CNVxplorer, to facilitate genetic diagnosis in a clinical setting.

## Materials and Methods

### High-confidence non-redundant sets of pathogenic and benign CNVs

Pathogenic and likely pathogenic CNVs were obtained from Clinvar (version 2021-10, [16]) and Decipher (version 2020-12-06, [17]), hereafter referred as the Clinvar and Decipher “pathogenic CNV sets”. Clinvar CNVs were restricted to those labelled as “copy number gain”, “copy number loss”, “deletion” or “duplication”, with clinical significance reported as pathogenic and germline origin, based on GRCh37 and with review status as “criteria provided, multiple submitters, no conflicts”, “criteria provided, single submitter” or “reviewed by expert panel”. Decipher pathogenic CNVs were restricted to *de novo* deletion or duplication variants, with a heterozygous genotype, labelled as “Pathogenic”, “Likely pathogenic” or “Unknown”, according to a previous study [15], and with contributions other than “none”. CNVs identified in the general population, hereafter collectively referred to as “benign CNV sets”, included those from the Decipher control set, the DGV database (version 2020-02-25), [18], gnomAD (controls-only dataset, version 2.1, [1]), and the Audano et al. [19], and Chaisson et Al. [20] sets. In the Decipher control set, CNVs with deletion_observations > 0 or (*exclusive or)* duplication_observations > 0 were retained. gnomAD variants were restricted to those with SVTYPE equal to “DEL” or “DUP” and FILTER equal to “PASS”. DGV variants were restricted to those with “varianttype” equal to “CNV” and “observedgains” > 0 or (*exclusive or)* “observedlosses” > 0. Only CNVs with “variantsubtype” equal to “deletion”, “duplication”, “loss” or “gain” were considered. The variants from Audano et al. and Chaisson et al. considered were restricted to those with a *remap* score equal to 1 and GRCh37.p13 assembly, whereas variants reported on a chromosome labeled as “unplaced” were filtered out. Variants from Beyter et al. [21] were mapped onto the GRCh37 human genome assembly with *liftOver* software [22], and were restricted to those of the same length not split into discontinuous genomic intervals after this process. For the DGV, gnomAD and Decipher control sets, only CNVs with an allele frequency no higher than 0.01 (1%) were considered, calculated as follows: (observed gains) / (sample size * 2) for duplications, and (observed losses) / (sample size * 2) for deletions. No allele frequency filtering was performed on the remaining benign CNV sets, because of a lack of information. CNVs of less than 50 bp in length or mapping to chromosome Y were also filtered out throughout this work.

We obtained non-redundant high-confidence CNV sets by applying the following filtering steps: Clinvar CNVs with the same genomic coordinates and conflicting pathogenicity annotations were removed. Decipher CNVs with the same genomic coordinates annotated as pathogenic, likely pathogenic, or unknown, or as benign, likely benign, or uncertain, were discarded. CNVs with at least a 30% overlap with problematic regions were removed. Problematic regions were obtained from the “Problematic Regions” UCSC track (https://genome.ucsc.edu/cgi-bin/hgTrackUi?db=hg19&g=problematic) including a set of 12 sources of genomic regions known to cause sequencing analysis artifacts. As a quality control, CNV coordinates from the GRCh37 (hg19) human genome assembly were mapped onto the GRCh38 (hg38) assembly with *liftOver* software [22], and only those that remained the same length, without splitting into discontinuous genomic intervals after this process were retained. The CNV coordinates and genomic annotations throughout this work refer to the GRCh37 assembly. For any pair of CNVs sharing a reciprocal overlap of at least 90% of their respective lengths based on genomic coordinates, we selected the shortest of the two. Filtering was then sequentially performed after sorting CNVs in descending order of the number of CNVs with which they presented reciprocal overlaps. Reciprocal overlap filtering was performed independently for deletions and duplications, and separately for the Clinvar pathogenic set, the Decipher pathogenic set and the benign set. After this process, pairs consisting of a pathogenic CNV from Clinvar or Decipher with a reciprocal overlap of at least 90% with a benign CNV were removed.

### CNV feature annotation

CNVs were annotated for 38 features (**Supplementary Table 2**) that could be grouped into two major categories, as follows:

#### Gene-based features

Genes for which at least 1 bp was targeted by the CNV genomic coordinates were extracted based on Ensembl Gene (version 103), using GRCh37.p13 reference human genome and annotated for the following gene-level features: (i) the probability of loss-of-function intolerance of the gene (pLI, version 2.1.1, [23]); (ii) the loss-of-function observed/expected upper bound fraction (LOEUF) score, version 2.1.1, [23]; (iii) the probability of being tolerant to both heterozygous and homozygous loss-of-function variants (pNull, version 2.1.1, [23]); (iv) the constrained coding region (CCR) score [24]; (v) the enhancer-domain score (EDS, [25]); (vi) prediction of haploinsufficiency or triplosensitivity, based on a meta-analysis of rare CNVs from 753,994 individuals [26]; (vii) ohnolog genes, as reported in the OHNOLOGS database (version 2, [27]) considering only pairs labeled as ‘strict’ (highly reliable); (viii) genes encoding transcription factors, according to data from the FANTOM consortium; (viii) fitness cost due to gene inactivation based on a genome-wide CRISPR-based score (CRISPR score, [28]), (ix) involvement of the proteins encoded by the genes in a protein complex (as extracted from hu.MAP 2.0, [29], with only proteins labelled with “extremely high” confidence selected); (x) mean and minimum gene expression across 54 tissues, obtained from the median transcripts per million (TPM) expression levels for each gene, available from https://storage.googleapis.com/gtex_analysis_v8/rna_seq_data/GTEx_Analysis_2017-06-05_v8_RNASeQCv1.1.9_gene_median_tpm.gct.gz as provided by the Genotype-Tissue Expression Project (GTEx; version 8, [30]) in which all GTEx tissues were considered); (xi) mean PhastCons 46-way placental score [31] and the CpG density of the promoter regions identified as 2 kb upstream and downstream from the transcription start site (TSS), defined as the first nucleotide of the transcript, according to previous work [32]. Transcript boundaries were obtained with BioMart from Ensembl Gene (version 103) [33], based on the GRCh37.p13 reference human genome. CpG density was calculated as follows:

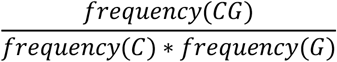

The sequence of each promoter was obtained with the R package BSgenome.Hsapiens.UCSC.hg19. (xii) Six network-based gene/protein features were extracted from a protein-protein interaction network: degree, PageRank, and shortest path to proteins associated with haploinsufficient and triplosensitive genes, respectively. For the construction of an unbiased network, protein-protein interactions were obtained from the STRING database (version 11.5, [34] available from https://stringdb-static.org/download/protein.links.full.v11.5/9606.protein.links.full.v11.5.txt.gz). Combined scores for edges were recalculated using only edges based on four types of evidence (presence of fusion, neighborhood, co-occurrence, and co-expression). The calculation of the combined score was based on a Python script (https://stringdb-static.org/download/combine_subscores.py) provided by the authors. Only high-confidence STRING interactions (>700 combined score) were finally retained. Gene-based features were transformed into categorical variables(“0” or “1” coding for the absence/presence, respectively, of at least one gene with the corresponding categorical feature), or quantitative variables encoding the maximum or minimum of the corresponding feature across the genes mapping within the CNV, except for minimum expression, the shortest path to haploinsufficient genes, and the shortest path to triplosensitive genes, for which the minimum was applied (**Supplementary Table 2**). In addition, missing values for the shortest path to haploinsufficient or triplosensitive genes were replaced by 4, the median estimate across all genes represented in the network.

#### Region-based features

The following region-based features associated with a given CNV were assessed: (i) the percentage of the CNV covered by each of the following six types of regulatory regions: open chromatin regions, transcription factor binding sites (TFBS), promoters, promoter flanking regions, CTCF sites, and enhancers, identified on the H1 human embryonic stem cell line (H1-hESC) and obtained from the Ensembl Regulatory Build (version 2019-11-01, [35]). (ii) The maximum recombination rate [36], CADD (version 1.6, [37]) and GERP scores [38] across the CNV genomic interval, with the scores previously summarized by their maximum value within non-overlapping 100 bp sliding windows. (iii) Maximum gene density across the 1 Mb sliding windows overlapping the CNV genomic interval. For the calculation of gene density, the human genome was split into non-overlapping 1 Mb sliding windows and the number of protein-coding genes mapping to each window was determined. As the last window of each chromosome could be different from 1 Mb, the final chunks were disregarded and 1 Mb windows were considered taking the end of the chromosome as the starting point. (iv) In addition, four features encoded the presence or absence of CNV overlap (i.e. at least 1 bp overlap) with the following regions of biological interest: (a) human accelerated regions (HARs), [39], (b) lamina-associated domains (LADs), [40], (c) ultra-conserved non-coding elements (UCNEs), [41]) and (d) structural variant (SV) hotspot regions [42]. (v) Two features encoded the distance (in megabases, Mb) to the centromere and to the closest telomere regions, with genomic coordinates retrieved from the track name “Gap” from UCSC Genome Browser [22]. Throughout this work, the minimum length required to consider a CNV to overlap a protein-coding gene or a genomic region was set at 1 bp.

### Training and testing CNV subsets

We sampled the following sets from the high-confidence non-redundant sets described above: a training CNV set built from Clinvar pathogenic CNVs evaluated before the year 2021, and benign CNVs; and two independent test sets built from (a) the Clinvar pathogenic CNVs evaluated since 1^st^ January 2021, and the benign CNV set; and (b) the Decipher pathogenic set and the benign CNV set. In these sets, pathogenic and benign CNVs were matched by length as follows: CNVs were assigned to discrete 100 bp length bins, ranging from 50 bp to 10^7^ bp. The same numbers of pathogenic CNVs and benign CNVs were retained in each bin, ultimately resulting in a 1:1 ratio within each non-empty bin. Within each bin, benign CNVs were prioritized, depending on their source, as follows: Beyter et al., Audano et al., Chaisson et al., gnomad_v2.1, DGV, and Decipher Control. This resulted in prioritization according to the underlying CNV detection technologies, i.e. long-read sequencing to short-read sequencing, to array comparative genomic hybridization (aCGH). For simplicity, these three matched-by-length sets of pathogenic and benign CNVs are referred to in the text as the Clinvar, Clinvar >2021, and Decipher sets. The Decipher set was as also considered as a training set, as an alternative to Clinvar, for the scoring of CNVs from the BANCCO database (http://bancco.fr).

We also constructed four challenging test scenarios by sampling CNVs from the Clinvar and Decipher pathogenic set, considered independent, fulfilling specific requirements: (i) Scenario #1, in which only pathogenic and benign CNVs targeting Mendelian disease genes were considered, as described below. (i) Scenario #2, in which only pathogenic and benign CNVs not targeting Mendelian disease genes were considered, as described below. (ii) Scenario #3, in which pathogenic CNVs not targeting Mendelian disease genes and benign CNVs targeting Mendelian disease genes were considered. (iii) Scenario #4, in which pathogenic and benign CNVs not targeting protein-coding genes were considered, as described below. In all these test sets, pathogenic and benign CNVs were obtained from the high-confidence and non-redundant sets described above and matched by length in a 1:1 ratio, as described above. We extracted a list of *n*=3762 Mendelian disease genes from the Online Mendelian Inheritance in Man (OMIM) database (version 2020-03-12;[43]). As described by Caron et al. [15], OMIM genes were restricted to monogenic Mendelian disease genes by filtering out genes associated with phenotype descriptions flagged as “somatic” or “complex” and with a supporting evidence level of 3 (i.e. the molecular basis of the disorder is known). We retrieved a list of *n*=18,792 protein-coding genes from the HUGO Gene Nomenclature Committee (HGNC, version 2021–03-03, [44]).

### CNVscore model implementation

CNVscore was implemented as the combination of a gradient tree-boosting model [11] and a Bayesian logistic regression model. Gradient-boosting was first trained to minimize the loss function *Log-loss* when classifying CNVs as pathogenic and benign on the training set and the *n*=38 features described above, without prior standardization or normalization. Missing data were imputed for the shortest path to a haploinsufficient gene and shortest path to a triplosensitive gene features. A value of 4 was imputed, as this was the median value across all genes for both features. Gradient boosting was trained with the R package rtemis (version 0.8.1). The loss minimization process across gradient-boosting training can be formally summarized as follows:

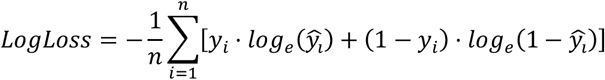

where *n* is the number of observations in the training dataset, 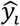is the predicted probability of the CNV being pathogenic, *y*_*i*_ is 1 if the CNV is pathogenic and 0 if the CNV is benign.

Potential contamination issues between training and testing CNVs in downstream applications were prevented by designing CNVscore as a bundle of 23 independent models each trained in a leave-one-chromosome-out manner across the autosomes and chromosome X. The scoring of a CNV in a given chromosome was thus calculated by the model that excluded the chromosome concerned from its training. Two independent bundles were trained on deletions and duplications, respectively. Hyperparameter selection for gradient-boosting training was performed by 10-fold cross-validation (AUROC as the evaluation metric) on the Clinvar training set across a total of 100 settings, each defined by a different combination of hyperparameters from the following values: number of trees ⊂[100, 200, 300, 400, 500], maximum depth ⊂[1, 2, 3, 4, 5], and minimum number of observations per node ⊂[1, 2, 3, 4]. The setting retained was that with 500 trees, a maximum depth of 2, and a minimum of 1 observation per node, which optimized performance for both deletions and duplications. In addition, a learning rate of 0.001 was selected. CNVscore training on the Decipher set was performed with the same hyperparameters as for the Clinvar set.

After the training process, each of the resulting trees was decoupled into a set of independent rules, as previously described [45]. These rules were used to classify the CNVs in the training set as pathogenic (1) or benign (0). Each CNV in the training set was then attributed a binary vector of 1s and 0s of dimensions equal to the number of rules extracted from the gradient-boosting trees. These vectors were then regarded as feature vectors and were used as input to train a Bayesian generalized linear regression model on the same training set in a leave-one-chromosome-out manner. Bayesian models were trained with the R package rstanarm (version 2.21.1). As a prior distribution, we used the regularized horseshoe prior for all coefficients. This prior is a global-local shrinkage prior characterized by a high concentration around zero [46], making it possible to perform pseudo-feature selection by shrinking small coefficients to close to zero. However, unlike the Laplacian distribution, the regularized horseshoe prior has heavy tails to prevent the excessive shrinkage of large coefficients [47]. The Bayesian models used the Markov Chain Monte Carlo (MCMC) algorithm as the posterior probability estimation approach. The mathematical formulation of the Bayesian regression was as follows:

#### Link function

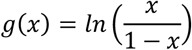

where *x* is the probability that *y* is equal to 1 (i.e. pathogenic).

#### Likelihood function

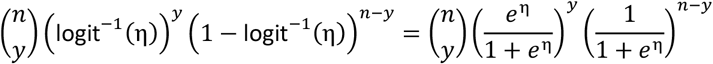

where *n* is the number of training observations, *y* is the number of pathogenic observations and η is a linear predictor with values in *R*.

#### Posterior distribution

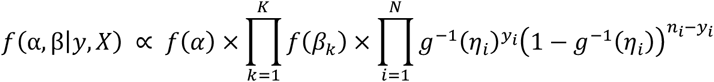

where *k* represents a coefficient and*K* is the total number of predictors, and *f*(*α*) and *f*(*β*_*k*_) represent the prior distribution of the intercept and predictors, respectively.

We evaluated two statistics to identify convergence problems of the MCMC sampler: effective sample size (ESS) and the potential scale reduction statistic (Rhat). Estimates were considered “unstable” if ESS < 1000 [48] or Rhat ≥ 1.1 [49]. With 3000 iterations, less than 25% of the estimates were considered to be “unstable”, across all Bayesian models throughout this work.

### Pathogenicity and uncertainty CNVscores

The Bayesian model described provided a posterior probability distribution (ranging between 0 and 1) for each CNV evaluated, indicating the probability of it being pathogenic. The posterior distributions for each CNV were then summarized with the median 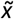 to assess centrality, and the standard deviation (*sd*) to assess dispersion. The 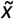 statistic was taken as the *pathogenicity* CNVscore for each CNV, whereas the *sd* values conveyed the uncertainty of the predictions. In addition, these two statistics were decorrelated by fitting a generalized additive model (GAM) between the *sd* and the pathogenicity CNVscore (i.e. formula “sd ∼ pathogenicity CNVscore”) with the R package mgvc (version 1.8.35) on the training CNVs. A Gaussian distribution (“gaussian”) and cubic regression splines (“cr”) were selected as family distribution and smooth term parameters, respectively. The residuals of each CNV in the training set were then quantile-normalized with respect to their 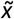 values as follows: first, the pairs of (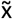, residual) values associated with the *n* CNVs in the training set were used to obtain *k* equal-sized bins across 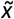 values, each containing ∼*n*/*k* CNVs. Within each bin, CNVs were ranked in ascending order, from lowest to highest residual values. Within each bin, ranks were transformed into rank-percentiles (ranging between 1 and 100). The smallest rank-percentiles of the residual were considered to correspond to the most reliable *pathogenicity CNVscore* prediction. CNVs with rank-percentiles within the [0-33], (33-66] and (66-100] ranges were considered to have low-, moderate- and high-uncertainty *CNVscore* levels, respectively, associated with a given *pathogenicity CNVscore* within the training set. The same procedure was performed independently on duplication CNVs from the training set. *k*=10 was set for a model trained on Clinvar deletions (*n*= 7486), and *k*=3 for Clinvar duplications and Decipher deletions and duplications, due to the smaller sample size: *n* = 392, 636 and 262, respectively (**Supplementary Table 3**).

Downstream scoring of query CNVs not used in the training set was performed by applying the corresponding gradient boosting-derived rules and Bayesian model to obtain the *pathogenicity* CNVscores and *sd* values. The raw *sd* value of each query CNV was then independently quantile-normalized by assignment to the 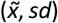 reference bins derived from the training set, as described above. Thus, query CNVs were assigned to the training-set 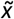 bin and uncertainty level (i.e. low, moderate or high) with the closest mid-point 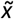 and *sd* values, respectively. Again, deletion and duplication CNVs were treated independently for scoring purposes. Experimental *p*-values estimating the statistical significance of the AUROC performance differences between the low- and high-uncertainty CNV subsets were estimated by bootstrapping. Thus, 80% of CNVs within each set were randomly drawn with replacement 100 times, with independent sampling of pathogenic and benign CNVs from each set. A two-sided Wilcoxon rank-sum test was performed on the two lists of *n*=100 AUROC values associated with the low- and high-uncertainty CNV subsets, respectively.

### CNV scoring methods considered in comparative benchmark tests

Benchmark analyses were performed with a set of five alternative CNV scoring methods representing the current state of the art: ClassifyCNV (version 1.1.1, [4]), CADD-SV (version 1, [8]), TADA (version 10 May 2021, [7]), STRVCTVRE (version 1.6, [5]) and X-CNV (version 23 Aug 2021, [6]). CNV genomic coordinates were converted from hg19 to hg38 coordinates with UCSC *liftOver* for use as input for CADD-SV and STRVCTVRE. All methods were run with default parameters. The main characteristics of the different approaches are described in **Table 1**.

### API implementation and integration into CNVxplorer

Interoperability with CNVscore was facilitated by implementing an application programming interface (API) supporting remote queries with the R package plumber (version 1.1.0). This API service can also be deployed as a private server through a Docker image without external dependencies, provided at https://github.com/RausellLab/CNVscore. Furthermore, CNVscore is integrated into the CNVxplorer webserver at http://cnvxplorer.com [3].

## Data Availability

All data produced in the present work are contained in the manuscript

https://github.com/RausellLab/CNVscore

## Acknowledgments

This study made use of data generated by the Decipher community. A full list of centers contributing to data generation is available from https://decipher.sanger.ac.uk/about/stats and via email from. Funding for the Decipher project was provided by Wellcome. BANCCO, the French National Database of Constitutional CNVs, is funded by the “Fondation pour la Recherche Médicale”. It is developed within the framework of the project “National NGS / ACPA databases” directed by Prof. Damien Sanlaville with the participation of Poitiers University Hospital (Dr. Frédéric Bilan), the University of Aix Marseille (Prof. Christophe Béroud) and the AChro-Puce network (https://acpa-achropuce.com). A full list of the centers contributing to data generation is available from https://bancco.fr/statistique.

## Supplementary Figures

**Supplementary Figure 1.**
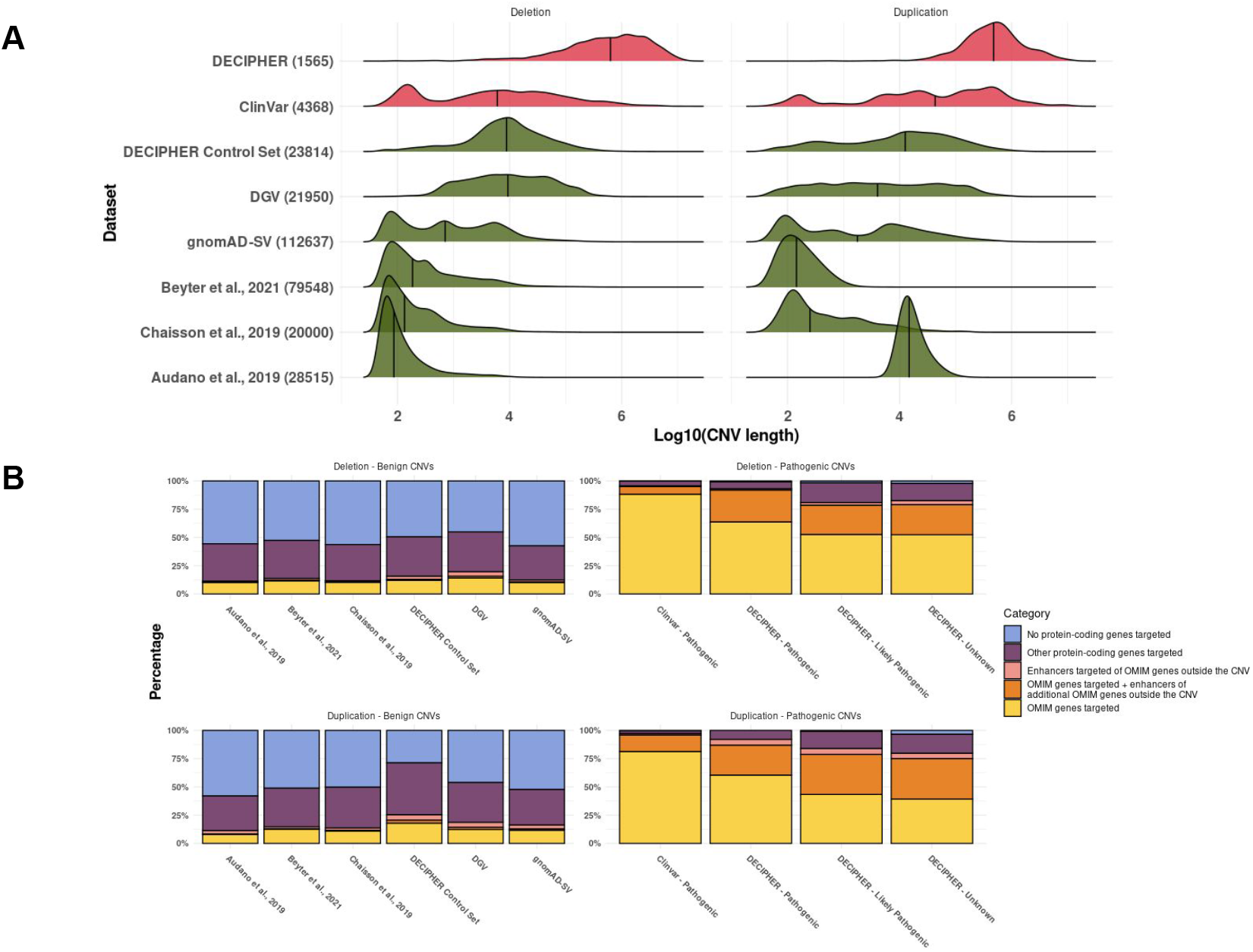
Genomic-length distribution and gene functional elements affected by reference pathogenic and benign CNV sets split by variant type, i.e: deletions and duplications. (A) CNV genomic length distributions (log-10 transformation of the number of base pairs) across pathogenic (red) and non-pathogenic (green) high-confidence non-redundant reference CNV sets (**Methods**). Median values are indicated with a vertical bar. The total number of CNVs within each set, for deletions and duplications separately, is indicated in parentheses. (B) Bar plots showing the percentage of CNVs within each reference set belonging to five different categories, according to the type of functional elements targeted. Categories were considered mutually exclusive and included the following: (i) CNVs targeting Mendelian disease genes (referred to in the legend as “OMIM” genes for simplicity; yellow); (ii) CNVs targeting Mendelian disease genes and enhancers of other Mendelian disease genes mapping outside the CNV boundaries (orange); (iii) CNVs with no direct impact on Mendelian disease genes, but with an indirect impact through enhancers of Mendelian disease genes (pink); (iv) CNVs with no impact on Mendelian disease genes, either direct or through enhancers, but with an impact on other protein-coding exons (dark violet); (v) CNVs with no impact on Mendelian disease genes, either direct or through enhancers, and no impact on any other protein-coding exons (light violet). CNV sets were grouped according to their classification as pathogenic (left) or benign (right; **Methods**).

**Supplementary Figure 2.**
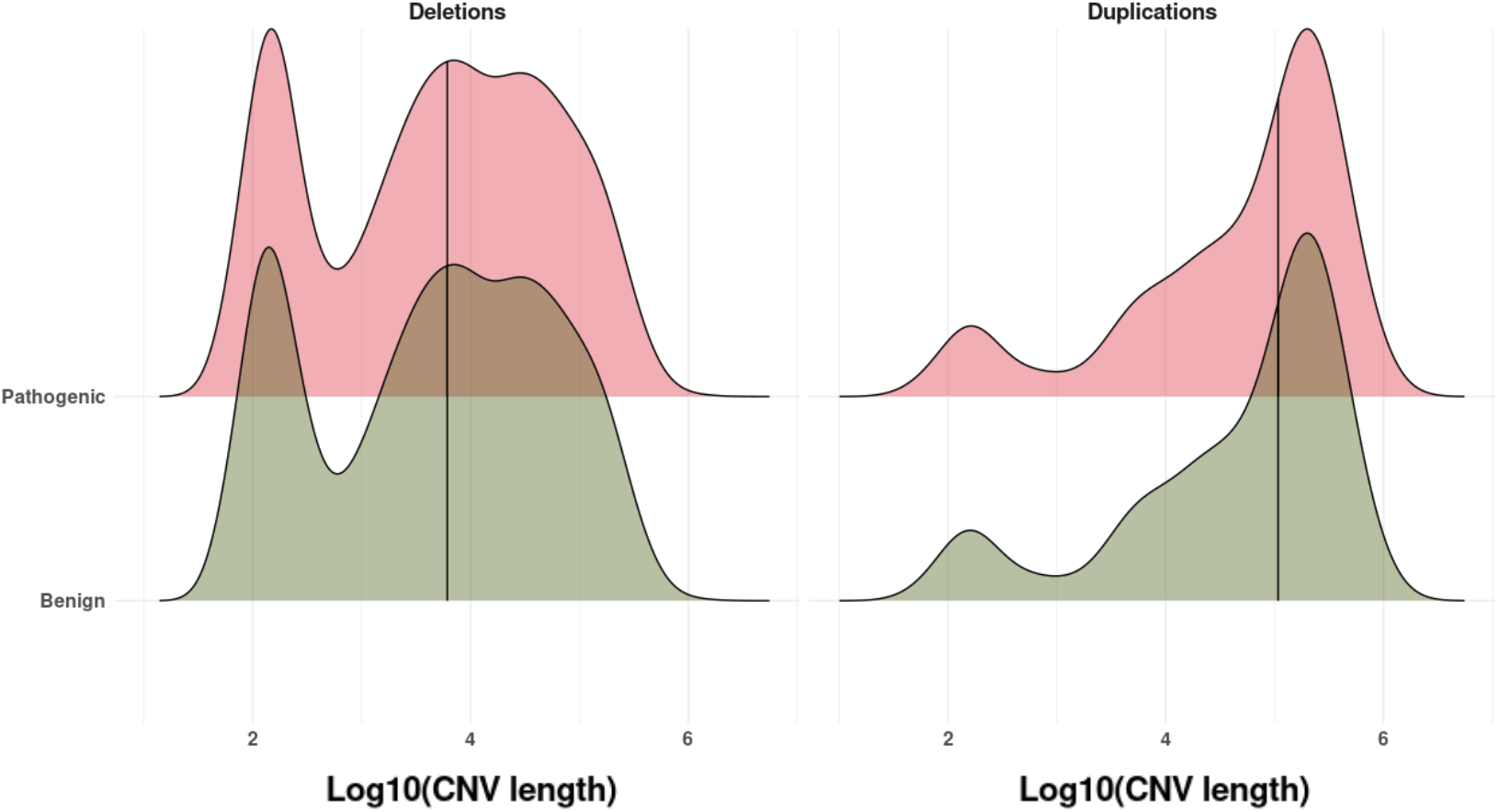
Length distribution of pathogenic and benign CNVs, split into deletions and duplications, of the Clinvar training dataset after matching by length. The CNV genomic length distribution (log-10 transformation of the number of base pairs) across pathogenic (red) and benign (green) high-confidence non-redundant reference CNV sets (**Methods**). Median values are indicated with a vertical bar.

**Supplementary Figure 3.**
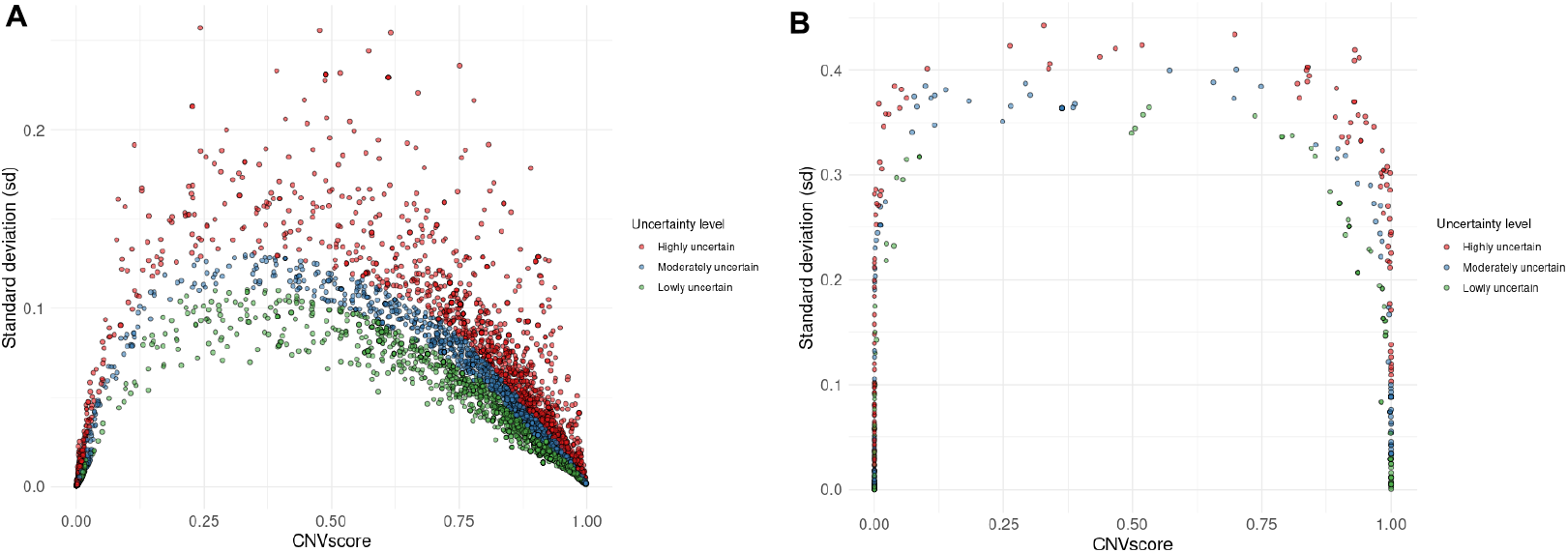
Uncertainty levels according to sd and pathogenicity CNVscore values for the Clinvar training dataset. This figure shows the pathogenicity CNVscores (*x*-axis) and standard deviation (sd) values (*y*-axis) associated with each deletion (A) and duplication (B), represented as dots, for the Clinvar training dataset (**Methods**). CNVs are colored according to their uncertainty level: low (green), moderate (blue) and high (red), calculated as described in the **Methods**.

**Supplementary Figure 4.**
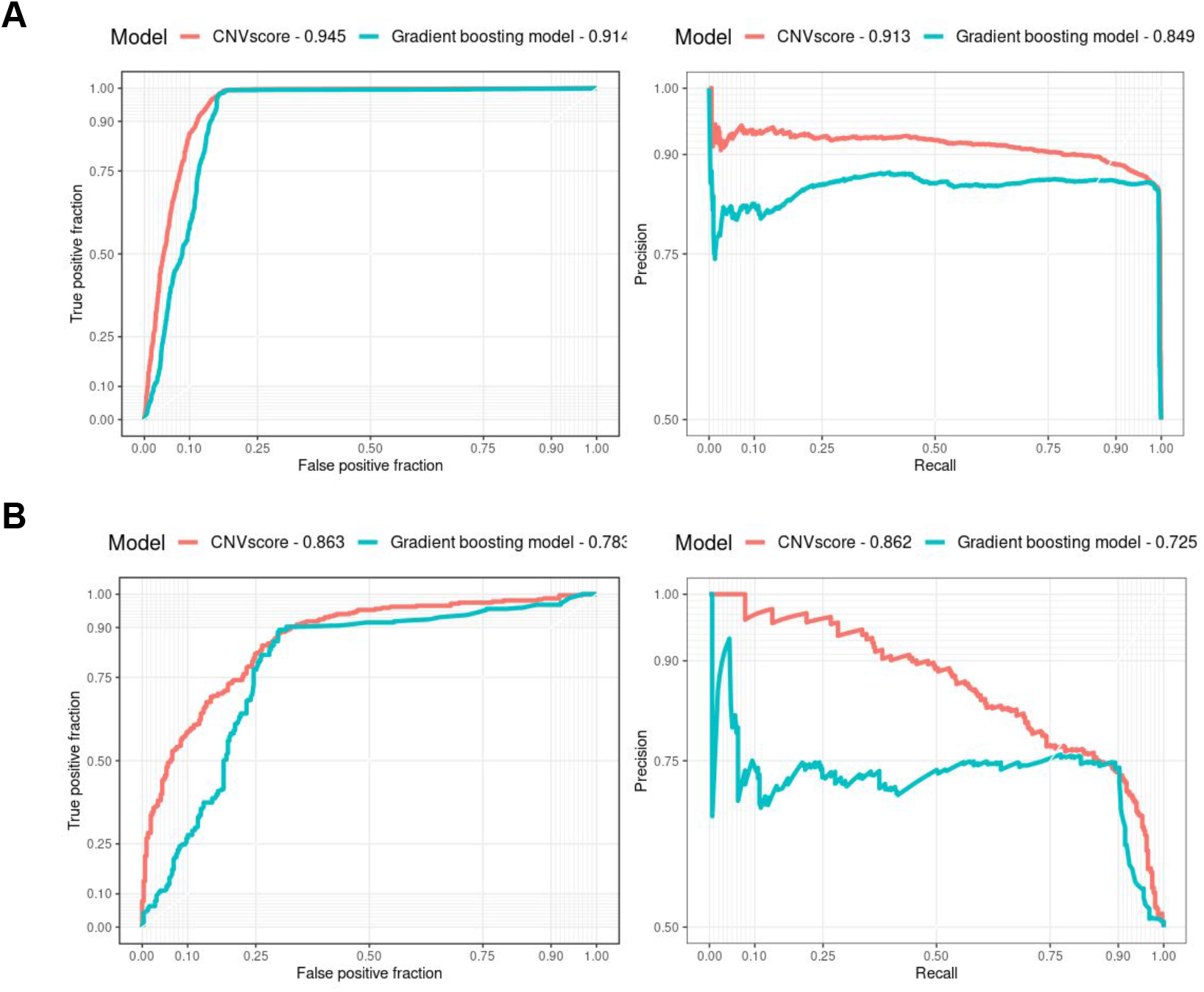
Performance comparison between a gradient-boosting model and CNVscore. This figure shows the ROC and PR curves of the gradient-boosting model (blue line) and CNVscore (red line) and their respective AUROC and AUPR values for the Clinvar (A) and Decipher (B) datasets.

